# “One more tool in the tool belt”: A qualitative interview study investigating patient and clinician opinions on the integration of psychometrics into routine testing for disorders of gut-brain interaction

**DOI:** 10.1101/2023.06.06.23291063

**Authors:** Mikaela Law, Esme Bartlett, Gabrielle Sebaratnam, Isabella Pickering, Katie Simpson, Celia Keane, Charlotte Daker, Armen Gharibans, Greg O’Grady, Christopher N. Andrews, Stefan Calder

**Author notes:** Corresponding Author: Dr Stefan Calder, Department of Surgery, The University of Auckland, Auckland, New Zealand. **Author Contributions:** The authors confirm their contribution to the paper as follows: study conception and design: ML, GS, CK, CD, AG, GOG, CNA, SC; data collection: ML, EB; analysis and interpretation of results: ML, EB, GS, IP, KS, CD, GOG, CNA, SC; draft manuscript preparation: ML, EB, GS. All authors reviewed, edited, and approved the final version of the manuscript.

## Abstract

**Background:** Psychological comorbidities are common in patients with disorders of gut-brain interaction (DGBIs) and are often linked with poorer patient outcomes. Likewise, extensive research has shown a bidirectional association between psychological factors and gastrointestinal symptoms, termed the gut-brain axis. Consequently, assessing and managing mental wellbeing, in an integrated care pathway, may lead to improvements in symptoms and quality of life for some patients. This study aimed to explore patients’ and gastroenterology clinicians’ opinions on integrating psychometrics into routine DGBI testing.

**Methods:** Semi-structured interviews were conducted with 16 patients with a gastroduodenal DGBI and 19 clinicians who see and treat these patients. Interviews were transcribed verbatim and analysed using inductive, reflexive thematic analysis.

**Results:** Three key clinician themes were developed: (1) psychology as part of holistic care, emphasising the importance of a multidisciplinary approach; (2) the value of psychometrics in clinical practice, highlighting their potential for screening and expanding management plans; and (3) navigating barriers to utilising psychometrics, addressing the need for standardisation and external handling to maintain the therapeutic relationship. Four key patient themes were also developed: (1) the utility of psychometrics in clinical care, reflecting the perceived benefits; (2) openness to psychological management, indicating patients’ willingness to explore psychological treatment options; (3) concerns with psychological integration, addressing potential stigma and fear of labelling; and (4) the significance of clinician factors, emphasising the importance of clinician bedside manner, knowledge, and collaboration.

**Conclusions:** The themes generated from the interviews indicated that patients and clinicians see value in integrating psychometrics into routine DGBI testing. Despite potential barriers, psychometrics would advance the understanding of a patient’s condition and facilitate holistic and multidisciplinary management. Recommendations for navigating challenges were provided, and considering these, patients and clinicians supported the use of psychometrics as mental health screening tools for patients with gastroduodenal DGBIs.

## Introduction

Gastroduodenal disorders of gut-brain interaction (DGBIs), formerly known as functional gastroduodenal disorders (FGDDs), are a significant global health concern due to their rising prevalence. At least 20% of the global population is estimated to meet the diagnostic criteria for a gastroduodenal DGBI, highlighting the need for effective diagnostic and management strategies [1–3]. These disorders include conditions such as functional dyspepsia (FD), gastroparesis (GP), and chronic nausea and vomiting syndrome (CNVS). The management and diagnosis of these disorders are complex as many patients present with gastroduodenal symptoms that lack an identifiable organic cause [4]. As a result, clinicians primarily rely on symptom classifications for diagnosis, resulting in overlapping etiologies due to shared symptomatology. Consequently, the availability of clear diagnostic pathways and targeted patient management has been limited.

A growing body of evidence has also demonstrated a bidirectional relationship between gastrointestinal symptoms and psychological factors in patients with gastroduodenal DGBIs (7–9). Psychological comorbidities are common in these patients, and stress, anxiety, and depression have been found to worsen symptoms and quality of life [5–11]. The gut-brain axis, a complex bidirectional neurohormonal pathway between the gastrointestinal tract and the brain, is key to understanding this association [5,12–14]. Psychosocial factors are also strong determinants for adherence, treatment effectiveness, functional ability, and healthcare utilisation and costs, further demonstrating the wide-ranging impact of psychological factors on the lives of patients with DGBIs [10,15]. Furthermore, psychological interventions and neuromodulators have been found to improve mental wellbeing and gastrointestinal symptoms in these patients [11,16–18]. The increasing recognition of this gut-brain axis and the adoption of a biopsychosocial framework has resulted in the integration of psychological approaches into the management of DGBIs as the recommended standard of care [2,5,10,19].

Therefore, early identification and management of psychological concerns may improve symptoms and reduce the need for extensive medical testing and trial-and-error treatments. This may have important implications for patients’ prognosis and perceived ability to cope with their symptoms. Accordingly, validating a method accepted by both patients and clinicians to identify those who would benefit from psychological support is crucial. Routine mental health screening is recommended in patients with DGBIs [5,10,15,20,21]. However, it is unclear how often this is done and how the results are utilised in clinical practice. Therefore, standardising mental health screening through psychometrics may provide an objective method of assessment. Incorporating psychometrics as part of routine clinical testing alongside physiological tests, such as body surface gastric mapping [22], could aid in the diagnosis and management of patients with gastroduodenal DGBIs and ultimately improve clinical care.

However, the need for psychometrics and how they would be used in practice is still unknown. This study aimed to investigate patient and clinician user needs and opinions on the integration of psychometrics within routine clinical testing for patients with gastroduodenal DGBIs, including whether psychometrics should be assessed, the advantages and disadvantages of this, and how the results should be included and utilised in clinical practice.

## Materials and Methods

### Design

Online semi-structured qualitative interviews were conducted with gastroenterology clinicians and patients with gastroduodenal DGBIs. Ethics approval was granted by the Auckland Health Research Ethics Committee (AH24466). The study was reported as per the Standards for Reporting Qualitative Research (SRQR) [23].

### Researcher Reflexivity

The researchers were a multidisciplinary group of health psychologists, medical professionals, and biomedical engineers specialising in gastric function, with diverse viewpoints. Some of the researchers had prior professional relationships with several of the clinician interviewees. This aided recruitment; however, it may have introduced bias, as these participants may have provided more positive feedback on the bases of these prior relationships. Interviews were conducted by a health psychology researcher (ML), who had no prior relationships with any of the interviewees. ML and EB conducted the analysis. ML has a broad background in qualitative and quantitative health psychology research, whilst EB was a third-year psychology student, interning as a health psychology researcher. The team’s perspectives and experiences informed the interview direction and data interpretation.

### Sample

Patients with FD, CNVS, or GP were recruited along with gastroenterology clinicians who commonly see and treat these patients. Patients were recruited via social media advertising and clinicians were approached directly via email. Inclusion criteria for both groups included being over the age of 18 and being able to speak and read English fluently. In addition, patients were eligible if they met the diagnostic criteria for CNVS and/or FD (as per the Rome IV criteria [24]), or had a diagnosis of gastroparesis, as confirmed by a standardised gastric emptying scan [25]. Patients were excluded if they had an eating disorder or self-induced vomiting. Interviews were concluded after 16 patients and 19 clinicians, as no new information was explored within the later interviews for each group, indicating thematic saturation. Recruitment and data collection were completed between July 2022 and February 2023.

### Protocol

Individual semi-structured interviews were conducted with each participant online via a video conferencing platform. Written informed consent and demographics were obtained prior to the interview. Separate interview schedules were created for patients and clinicians. Guiding questions were developed based on the study’s aims, a scoping survey with clinicians, and in consultation with two gastroenterologists (CNA and CD) who ensured question clarity and ease of understanding. The interview schedules were used to guide the discussion, but the semi-structured style allowed for flexibility to explore any topics that arose.

Patient interviews explored their experiences of being asked about psychological information by their gastroenterology clinician(s) and their views on integrating psychometrics into routine testing for their condition. This included the perceived usefulness of psychometrics, how they would want their clinician to use the results, their level of comfort discussing these results with clinicians, and anticipated problems or challenges. Similarly, clinician interviews explored their existing approaches for psychological assessments in these patients, as well as their views on the integration of psychometrics into routine testing. This included their perception of the usefulness of psychometrics and its influence on clinical practice, how they would utilise the results, their level of comfort using this data, and any anticipated problems or challenges. All interviews were audio-recorded and transcribed verbatim, with both clinicians and patients given the option to read and correct their transcriptions before analysis.

### Data Analysis

Inductive, reflexive thematic analysis was conducted by two coders (ML and EB) [26,27]. The use of two coders allowed for a collaborative approach, which enhanced reflexivity and triangulation as the themes were developed through two diverse, subjective lenses. The coders initially read the transcripts to familiarise themselves with the data set and independently began generating codes of information from the transcripts. These codes were then iteratively grouped into common patterns of meaning (themes). The two coders then began a collaborative process of discussing, defining, and refining the developed themes. These themes were refined collaboratively until a narrative was formed based on a set of key themes and subthemes developed from the analytic process. This process was done separately for the clinician and patient data sets to generate two sets of results.

## Results

### Demographics

A comprehensive overview of the demographic characteristics of the sample is presented in *Table 1*. In total, 16 patients and 19 clinicians were interviewed. As shown in *Table 1*, all patients met the Rome IV criteria for FD, with 13 also meeting the criteria for either CNVS and/or GP. The clinician sample comprised both physicians and surgeons, specialising in various areas of gastroenterology, who regularly see and treat patients with gastroduodenal DGBIs. Four clinicians were currently completing a residency or fellowship. The patient interviews lasted on average 39 minutes (range= 25-52 minutes) and the clinician interviews lasted on average 29 minutes (range= 19-37 minutes).

**Table 1.** Demographic and clinical characteristics of the sample

|  | Patients (n= 16) | Clinicians (n= 19) |
| --- | --- | --- |
| Age, M (range) | 34.88 (20-55) | 44.05 (30-67) |
| Gender, n (%) |  |  |
| Female | 14 (88%) | 5 (26%) |
| Male | 2 (12%) | 14 (74%) |
| Ethnicity, n (%) |  |  |
| White/Caucasian | 10 (63%) | 11 (58%) |
| Asian | 2 (12%) | 6 (32%) |
| Other | 3 (18%) | 1 (5%) |
| Not stated | 1 (6%) | 1 (5%) |
| Country of residence, n (%) |  |  |
| New Zealand | 6 (38%) | 9 (47%) |
| Australia | 5 (31%) | 2 (11%) |
| Canada | 3 (19%) | 5 (26%) |
| United States | 1 (6%) | 2 (11%) |
| Scotland | 1 (6%) | 0 (0%) |
| Belgium | 0 (0%) | 1 (5%) |
| Diagnosis, n (%) |  |  |
| FD, CNVS, and GP | 10 (63%) | - |
| FD and CNVS | 2 (12%) | - |
| FD and GP | 1 (6%) | - |
| FD only | 3 (19%) | - |
| Occupation, n (%) |  |  |
| Physician | - | 16 (84%) |
| Surgeon | - | 3 (16%) |
| Speciality, n (%) |  |  |
| General gastroenterology | - | 8 (42%) |
| Gastric motility | - | 8 (42%) |
| Other | - | 3 (16%) |
| Years of experience, M (range) | - | 10.26 (2-30) |

*Note.* M=mean; SD= standard deviation; %= percentage of participants in that category; CNVS= chronic nausea and vomiting syndrome; FD= functional dyspepsia; GP= gastroparesis

### Clinician Themes

Three key themes, each with a number of subthemes, were developed from the analysis of the clinician interviews. The hierarchy of themes is shown in *Table 2*, with example quotations, and described briefly in-text. Subthemes have been presented in-text using italics.

**Table 2.** Hierarchy of the themes and subthemes identified from the clinician interviews with example quotations.

| Key Theme | Subtheme | Quotes |
| --- | --- | --- |
| **1. Psychology as part of holistic care** | Awareness of gut-brain connection | "I think there is a strong connection between functional GI issues and mental health issues, especially depression and anxiety. And then certainly, stress is one of those all-encompassing things that can play a role in both exacerbating symptoms and also can be a barrier for improving things." (C01)<br>"I think with these types of patients we need to... take a look at them as a whole patient, rather than just the disease that we diagnose them with." (C19) |
|  | Relevance of gathering psychological information | "It's pretty routine that I would ask [about mental wellbeing], and even if I don't, patients will sometimes volunteer it... For me it's a pretty fundamental component of the history." (C12)<br>"Rather than just focusing on physical symptoms, it's [asking about mental wellbeing] to try and get a more global impression of the problems that might be at play." (C06) |
|  | Psychology as a component of multidisciplinary management | "I've read the literature about the cognitive-behavioural therapy... those [patients] that would be receptive to it, I think I would be totally comfortable in recommending it. Because I know it would probably work for them." (C04)<br>"If we had access to a multi-disciplinary team with expertise in dealing with patients with functional gut disorders which included a dietitian, a physiologist, a psychiatrist, a psychologist, your surgeon, your gastroenterologist, your everyone, that would be fabulous." (C07) |
| **2. Value of psychometrics in clinical practice** | Psychometrics not widely used | "I think validated questionnaires are so important, and some people I know use them, but I haven't used them during my motility training specifically, and I don't know many people that use them routinely for functional patients." (C01)<br>"One of the other systemic reasons would be the organisation required to actually get people to do them [psychometrics]... It'd have to be a kind of a formalised process, I think. But if that was able to be done pre-clinic, then I think it would be really useful." (C09) |
|  | Psychometrics as screening tools | "In a perfect world I would screen almost every single one of my patients for psychological issues and then be able to refer them to appropriate care." (C07)<br>"It'd be good to have a proper way of formally assessing what their mental status is like... I think if we had that there that would be really helpful." (C05) |
|  | Advancement of understanding | "I think it [psychometrics] would be helpful because that would provide some additional information about what you may want to focus on." (C01)<br>"I think it would open the door to a very important discussion with the patient with a little bit of |
|  |  | data that you can actually talk to them about.” (C08) |
|  | Allows for expansion of management plan | “[Psychometrics] may improve your efficiency a little bit, in the sense that you would be able to then target or prioritise a few things differently.” (C01)<br>“Use it [psychometrics] as a gateway to help refer them, or get that under control... We can only do so much from our standpoint or some medications... And so, all together, we have better chances of success.” (C15) |
| **3. Navigating barriers to utilising psychometrics** | Patient stigma | “There'll be a subgroup of patients that will choose to not provide it [psychometric data], because of the fact that they've been stigmatised in the past.” (C17)<br>“Trivialising it and making it as a routine part of the assessment... so it's not like you've just reacted to what they're telling you, that that you think they're crazy, rather than like this is a standard part of the workup. I think that sort of maybe takes away some of the stigma” (C02) |
|  | Clinician knowledge | “So the main issue is that the clinicians who are likely to be requesting the test are likely to be clinicians who don't have the skillset for dealing with the psychological results.” (C07) |
|  | Ability to act | “My impression is that mental health services are pretty stretched.” (C06)<br>“The problem is around access. Our psychologist is a very valuable and limited resource. And so generally, it's kind of the end of the spectrum ones who get the psychologist.” (C07) |
|  | Responsibility for data | “We're meant to, as professionals, have a responsibility to make sure that people are not going to hurt themselves or somebody else... I don't know where you'd have to draw the line. Like if they said that they were, for example, got 27/27, and were majorly depressed... and they jumped off the bridge. Would we be held liable knowing that they'd shared that information with us?” (C05)<br>“It then raises an important question... Who should have access to this information? Other health professionals? Can family members access this?” (C17) |
|  | Clinician burden | “I think overall the time would be less, because I think a lot of these patients there is massive psychological overlay, and you spend ages seeing them and following them up in clinic every six months. And maybe the issue is that they are really anxious or depressed.” (C05) |

#### Theme 1: Psychology as part of holistic care

The first clinician theme reflected clinicians’ views on the importance of psychology as part of providing holistic and integrated healthcare for patients with gastroduodenal DGBIs. Clinicians had an *awareness of the gut-brain connection* and the bidirectional association between gastrointestinal symptoms and psychological factors. This was evidenced by the fact that patients with gastroduodenal DGBIs often had comorbid mental health issues. They believed psychology is an essential component that was inseparable from the physical aspects of the condition, thereby emphasising the need to assess a patient’s mental wellbeing.

In line with this understanding, clinicians discussed the *relevance of gathering psychological information* in this patient group. All clinicians interviewed inquired about patients’ mental wellbeing during unstandardised conversations or informal history taking, with some making it routine practice, while others only if there is a cause for concern. Clinicians also mentioned that patients also often volunteered this information unprompted.

Gathering psychological information was seen as a useful way to incorporate *psychology as a component of multidisciplinary management,* therefore providing another treatment avenue for patients. Targeting both psychological and physiological treatment pathways was seen as critical for improving gastrointestinal symptoms and quality of life. Clinicians raised a desire for a multidisciplinary system with integrated psychological and gastroenterology services. However, currently, the two services were perceived to be “splintered” (C09), making integrated care difficult.

#### Theme 2: Value of psychometrics in clinical practice

The second clinician theme explored the perceived value of integrating psychometrics as part of routine testing for patients with gastroduodenal DGBIs. Firstly, clinicians acknowledged how formal, standardised *psychometrics were not widely* used in current clinical practice, due to limited appointment times and lack of knowledge on which questionnaires to use. Systemic reasons also play a role as psychometrics were not considered part of routine gastroenterology testing. Despite this, all clinicians recognised their value and desired their integration into standardised routine assessments for these patients, especially if done externally.

In particular, clinicians supported the use of standardised *psychometrics as screening tools* to identify potential mental health concerns, particularly for anxiety, depression, and stress. Although other psychological information was mentioned as important to assess (e.g. eating disorders, social support, personality disorders), it was determined that it would be most appropriate to first screen for these basic mental health concerns and then conduct more in-depth psychological explorations if needed. Although not diagnostic, this approach was perceived as less intrusive and more objective, standardised, and validated than current informal assessment methods.

Additionally, psychometrics were seen as valuable for the *advancement of understanding* of a patient’s condition. They would allow for the combination of physiological and psychological data, aiding a more holistic view of a patient’s health and wellbeing. This provides a deeper exploration into how the brain and gut may interact to produce, exacerbate, and maintain the patient’s symptoms. Conversely, psychometrics can be used to discount psychological factors as significant in a patient’s symptomology. Psychometrics can also be used as a tool to explain the gut-brain axis to a patient to increase their own understanding.

Lastly, psychometrics could *allow for the expansion of the management plan* to include psychological intervention pathways, alongside medical management. Although not diagnostic, psychometric screening could act as a prompt for both patients and clinicians to consider psychological referrals, behavioural interventions, and neuromodulators, leading to a more tailored and multidisciplinary treatment plan. This is particularly beneficial for a patient group with limited effective medical options as it may provide other pathways for support. Psychometrics may also encourage patients to manage their own mental health through psychoeducation, self-management, or digital self-help apps.

#### Theme 3: Navigating barriers to utilising psychometrics

The final clinician theme explored the potential barriers to utilising psychometrics within clinical practice and how these can be navigated. Firstly, clinicians discussed the presence of *patient stigma* around mental health. Many patients fear that their symptoms will be blamed on mental health and medical treatment withheld, making them hesitant to discuss their mental wellbeing. Awareness of this stigma makes it difficult for clinicians to broach this topic early on in the patient-clinician relationship. Consequently, clinicians usually waited until they had developed a rapport with their patients. However, they preferred to explore psychological factors earlier and communicated how external and standardised assessments would facilitate discussions about mental health while maintaining the therapeutic relationship. Clinicians also stressed the importance of prefacing a patient using patient-friendly language before administering a psychological questionnaire, to explain why the information is being collected, how the data will be used, and that they can opt-out.

*A lack of clinician knowledge* was also mentioned as gastroenterology clinicians may not have the skills or expertise to deal with psychological concerns. However, some clinicians stated that although they were not explicitly trained in this area, psychological issues are a key component of DGBIs and therefore they had learnt about psychology through their clinical practice. Others may require training on how to interpret and use the results to feel more comfortable utilising psychometrics in practice.

Clinicians were also concerned about their *ability to act* on the results of the psychometrics, as they are not trained in providing psychological treatments. Referral to psychological services was suggested, but availability and accessibility were a concern, particularly for health psychologists specialising in psychogastroenterology. This may result in long waiting times or an inability to access treatment. In such cases, self-management protocols or digital apps were seen as useful.

Clinicians also raised concerns over *responsibility for psychometric data*. There was concern clinicians may not be comfortable being responsible or liable for this data, especially for questions about suicide or self-harm, as gastroenterology clinicians may not have the capacity to act on these concerns immediately. An opt-in model was therefore preferred to allow clinicians to understand the responsibility and take ownership of the data. Additionally, clinicians were concerned about data privacy and access. They emphasised the need for a patient-friendly presentation of results and awareness of who could access the data and its potential repercussions.

Finally, it was mentioned that the integration of psychometrics could increase *clinician burden,* causing extra work in reviewing, discussing, and following up with patients. However, the majority of the clinicians believed that using psychometrics would be an overall time-saver, especially if this was conducted externally and not during clinic time.

### Patient Themes

Four key themes, with several subthemes, were developed from the patient interviews. The hierarchy of the themes is outlined in *Table 3* with example quotations and described in-text.

**Table 3.**
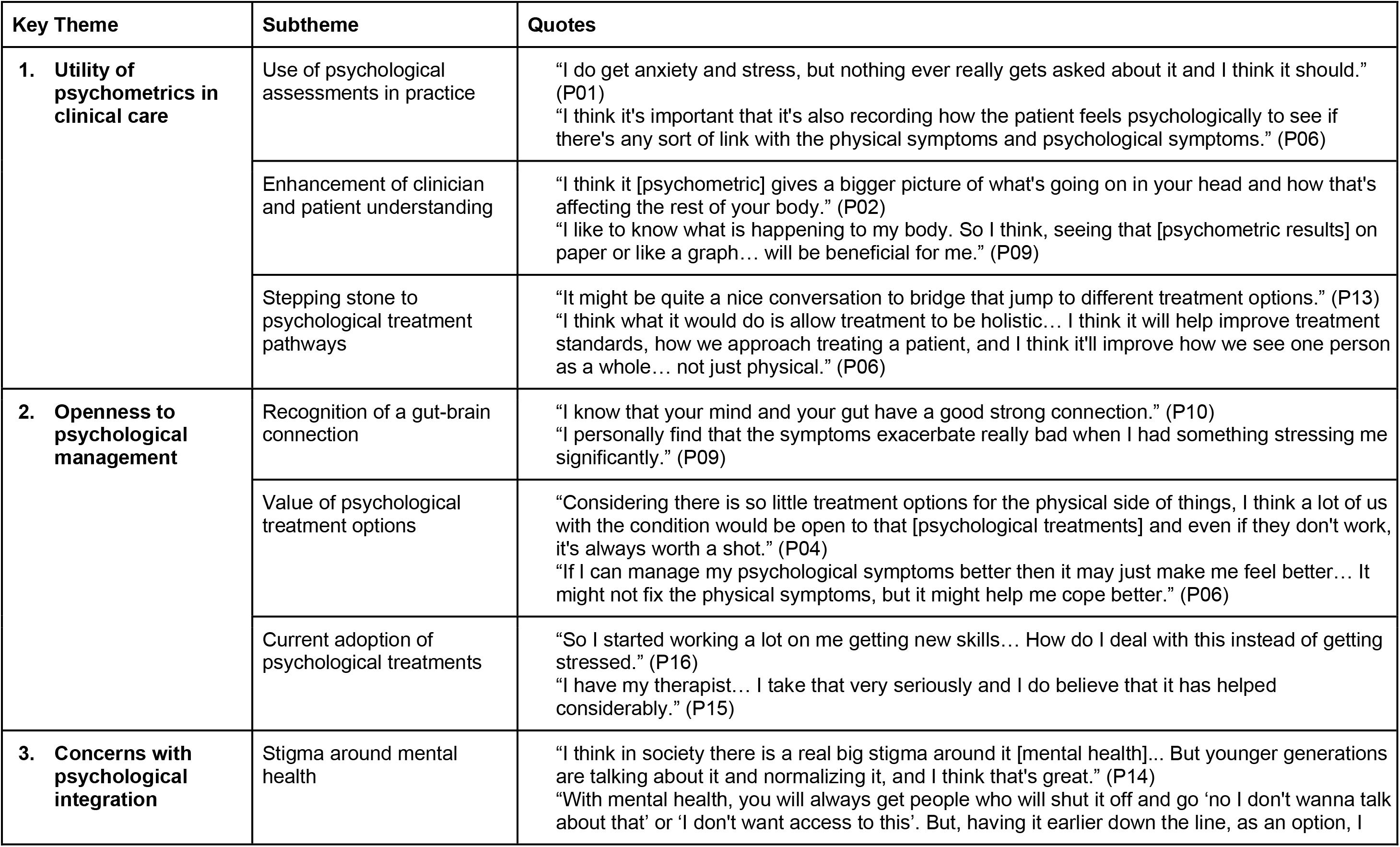
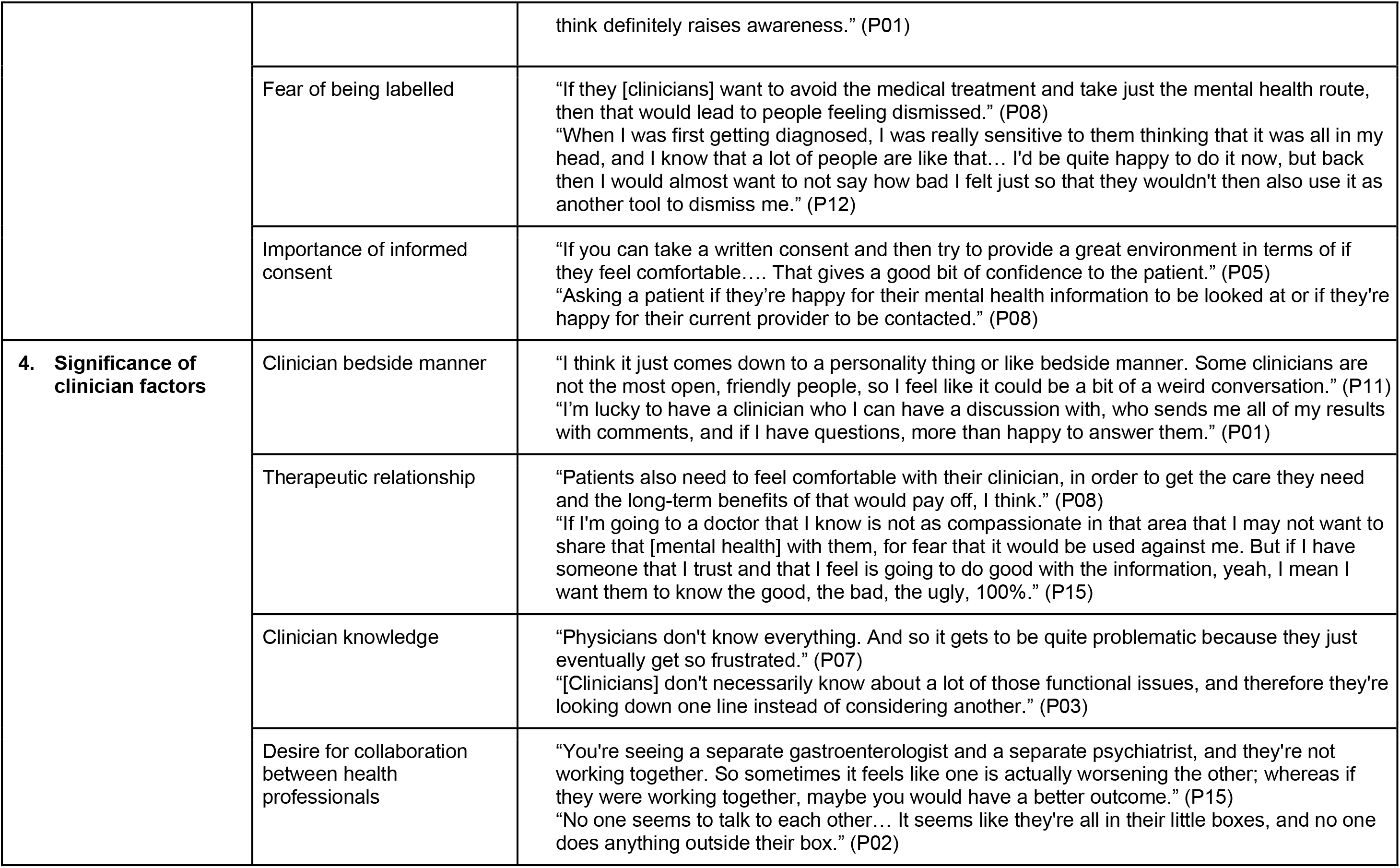
Hierarchy of the themes and subthemes identified from the patient interviews with example quotations.

| Key Theme | Subtheme | Quotes |
| --- | --- | --- |
| **1. Utility of psychometrics in clinical care** | Use of psychological assessments in practice | "I do get anxiety and stress, but nothing ever really gets asked about it and I think it should." (P01)<br>"I think it's important that it's also recording how the patient feels psychologically to see if there's any sort of link with the physical symptoms and psychological symptoms." (P06) |
|  | Enhancement of clinician and patient understanding | "I think it [psychometric] gives a bigger picture of what's going on in your head and how that's affecting the rest of your body." (P02)<br>"I like to know what is happening to my body. So I think, seeing that [psychometric results] on paper or like a graph... will be beneficial for me." (P09) |
|  | Stepping stone to psychological treatment pathways | "It might be quite a nice conversation to bridge that jump to different treatment options." (P13)<br>"I think what it would do is allow treatment to be holistic... I think it will help improve treatment standards, how we approach treating a patient, and I think it'll improve how we see one person as a whole... not just physical." (P06) |
| **2. Openness to psychological management** | Recognition of a gut-brain connection | "I know that your mind and your gut have a good strong connection." (P10)<br>"I personally find that the symptoms exacerbate really bad when I had something stressing me significantly." (P09) |
|  | Value of psychological treatment options | "Considering there is so little treatment options for the physical side of things, I think a lot of us with the condition would be open to that [psychological treatments] and even if they don't work, it's always worth a shot." (P04)<br>"If I can manage my psychological symptoms better then it may just make me feel better... It might not fix the physical symptoms, but it might help me cope better." (P06) |
|  | Current adoption of psychological treatments | "So I started working a lot on me getting new skills... How do I deal with this instead of getting stressed." (P16)<br>"I have my therapist... I take that very seriously and I do believe that it has helped considerably." (P15) |
| **3. Concerns with psychological integration** | Stigma around mental health | "I think in society there is a real big stigma around it [mental health]... But younger generations are talking about it and normalizing it, and I think that's great." (P14)<br>"With mental health, you will always get people who will shut it off and go 'no I don't wanna talk about that' or 'I don't want access to this'. But, having it earlier down the line, as an option, I |
|  |  | think definitely raises awareness.” (P01) |
|  | Fear of being labelled | “If they [clinicians] want to avoid the medical treatment and take just the mental health route, then that would lead to people feeling dismissed.” (P08)<br>“When I was first getting diagnosed, I was really sensitive to them thinking that it was all in my head, and I know that a lot of people are like that... I'd be quite happy to do it now, but back then I would almost want to not say how bad I felt just so that they wouldn't then also use it as another tool to dismiss me.” (P12) |
|  | Importance of informed consent | “If you can take a written consent and then try to provide a great environment in terms of if they feel comfortable.... That gives a good bit of confidence to the patient.” (P05)<br>“Asking a patient if they're happy for their mental health information to be looked at or if they're happy for their current provider to be contacted.” (P08) |
| **4. Significance of clinician factors** | Clinician bedside manner | “I think it just comes down to a personality thing or like bedside manner. Some clinicians are not the most open, friendly people, so I feel like it could be a bit of a weird conversation.” (P11)<br>“I'm lucky to have a clinician who I can have a discussion with, who sends me all of my results with comments, and if I have questions, more than happy to answer them.” (P01) |
|  | Therapeutic relationship | “Patients also need to feel comfortable with their clinician, in order to get the care they need and the long-term benefits of that would pay off, I think.” (P08)<br>“If I'm going to a doctor that I know is not as compassionate in that area that I may not want to share that [mental health] with them, for fear that it would be used against me. But if I have someone that I trust and that I feel is going to do good with the information, yeah, I mean I want them to know the good, the bad, the ugly, 100%.” (P15) |
|  | Clinician knowledge | “Physicians don't know everything. And so it gets to be quite problematic because they just eventually get so frustrated.” (P07)<br>“[Clinicians] don't necessarily know about a lot of those functional issues, and therefore they're looking down one line instead of considering another.” (P03) |
|  | Desire for collaboration between health professionals | “You're seeing a separate gastroenterologist and a separate psychiatrist, and they're not working together. So sometimes it feels like one is actually worsening the other; whereas if they were working together, maybe you would have a better outcome.” (P15)<br>“No one seems to talk to each other... It seems like they're all in their little boxes, and no one does anything outside their box.” (P02) |

#### Theme 1: Utility of Psychometrics in clinical care

The first key patient theme explored the perceived utility of psychometrics in clinical practice. Patients had mixed past experiences with *the use of psychological assessments in practice.* While some patients had been informally asked about their mental wellbeing by their gastroenterology clinicians, many patients stated their clinicians had never asked about their mental health. Often, these questions were only asked if a patient had a history of a mental health condition or if the clinician had exhausted other explanations for the patient’s symptoms. Patients highlighted how gastroenterology clinicians tended to only focus on the physical, emphasising a clear separation between the body and mind within their practice. None of the patients interviewed had ever been asked to complete a psychological questionnaire as part of their condition assessment. However, patients saw the value of integrating psychometrics into clinical care as part of a more holistic health assessment.

Patients saw the benefit of psychometrics for the *enhancement of clinician and patient understanding.* Combining physiological and psychological information would provide a bigger picture of how the brain and body interact and how this affects symptomology. Patients were eager to learn more about their condition and therefore desired to be informed about their psychometric results to further add to their own understanding of their symptoms.

Patients also explored how psychometrics could act as *a stepping stone to psychological treatment pathways.* Patients viewed psychometrics as an effective way to prompt the consideration of psychological referrals or interventions as part of a more integrated and tailored treatment plan. Psychometrics could also encourage patients to think about how self-management of their mental health could be beneficial.

#### Theme 2: Openness to psychological management

The second key patient theme demonstrated patients’ openness to psychological treatment options. Patients had a strong *recognition of a gut-brain connection* and thus understood the importance of managing their mental health in relation to their symptoms. This awareness evolved from personal experiences and research. They explored how this connection was bidirectional; psychological factors, such as stress or anxiety, can trigger or exacerbate gastrointestinal symptoms and, on the other hand, gastrointestinal symptoms can also lead to mental health issues.

Because of this knowledge, patients saw *the value of psychological treatment options,* especially if integrated as part of a multidisciplinary approach. This additional management option was seen as particularly useful as current medical treatments were generally perceived by patients as ineffective for symptom management. Patients discussed their mixed preferences for face-to-face or digital psychological interventions, highlighting the need for a choice of options.

In fact, many patients *currently adopt psychological treatments* as part of their symptom management, further demonstrating their openness. Some reported using self-management techniques (e.g. stress reduction, self-help apps), while others took psychiatric medications or regularly visited clinical psychologists. Overall, patients found these treatments helpful in improving both their mental wellbeing and gastrointestinal symptoms. These psychological techniques were frequently self-initiated and independent from the management plans provided by their gastric clinicians, further highlighting their desire to harness psychological treatments.

#### Theme 3: Concerns with psychological integration

The third key patient theme explored the concerns that patients had about the integration of psychology within their clinical care. Firstly, patients expressed concern about the potential *stigma around mental health*, which could discourage some patients from completing psychological assessments and treatments. However, the patients also discussed how it was becoming more acceptable to talk about mental health and the need to continue to normalise and spread awareness of the gut-brain axis.

Part of this stigma was considered to be due to the *fear of being labelled*. Patients were worried that the results of psychological assessments may be misinterpreted, leading to the clinician thinking that they are a “head case” (P02) or that their symptoms are “all in their head” (P16). This labelling may result in a patient not receiving proper medical care, something some of the patients interviewed had experienced in the past. Despite these concerns, the patients recognised the significance of psychological assessments and treatments as part of holistic management. Providing reassurance, where appropriate, that medical treatment will not be withheld could minimise these concerns.

Because of the above concerns, patients emphasised *the importance of informed consent* and highlighted the need for psychological assessments, referrals, and interventions to always be presented as optional. They also expressed the need for transparency regarding who had access to their data, as psychometrics collect sensitive information.

#### Theme 4: Significance of clinician factors

The final patient theme explored the influence of clinician factors on patients’ acceptance of psychological integration. Patients discussed the influence of *clinician bedside manner,* highlighting the importance of clinicians’ sensitive approach when discussing mental health. They listed clinician attributes that would make them feel more comfortable (e.g. open-mindedness, compassion, and empathy). Many felt lucky to have a clinician like this; although, they had also encountered clinicians who were less supportive. They would therefore prefer to search for clinicians with whom they felt more comfortable. However, they acknowledged that access to such clinicians may be limited, especially in rural areas.

Further, the *therapeutic relationship* was also perceived to be a critical determinant of patient comfort. Patients emphasised the importance of building trust and rapport with their clinicians so that they can be reassured their psychological data will not be misused. Communication was seen as crucial to building this therapeutic relationship and patients felt a sense of trust with clinicians that involved the patient in management decisions by being open to listening and receptive to the patient’s considerations. This mutual understanding allows patients to feel more comfortable discussing their mental health and exploring psychological treatment options.

Patients also explored the importance of *clinician knowledge* and discussed how they were more comfortable discussing their mental health with clinicians who were knowledgeable enough to use this information in a positive way. There was a perception that gastroenterology clinicians lack knowledge in psychology since their training primarily focused on the body. Patients stressed the significance of educating clinicians about DGBIs and their connection to mental health to avoid mislabelling, inappropriate treatments, or multiple referrals, and to facilitate the positive use of psychometrics.

Lastly, patients emphasised a *desire for collaboration between health professionals* to allow for multidisciplinary and integrative care. They discussed the current disconnect between clinicians, who often adhered to their own specialities, with little communication. In contrast, patients believed that a collaborative and multidisciplinary approach would greatly improve their management by leading to more tailored and integrative treatments, a normalisation of psychology, and an increase in the accessibility to psychologists.

## Discussion

This study employed semi-structured interviews to explore the user needs and opinions of clinicians and patients on the integration of psychometrics into routine clinical testing for gastroduodenal DGBIs. The themes generated from the interviews indicated agreement on the value of psychometric integration, despite potential concerns.

Both patients and clinicians recognised the importance of the gut-brain axis, a concept which has been gaining momentum within DGBI research [2,14,17,28]. This highlights the importance of examining mental health as part of an integrated assessment for patients with gastroduodenal DGBIs to improve understanding of their condition and screen for potential psychological comorbidities, which are common in this patient population [10,17]. Various clinician guidelines agree that mental wellbeing is important to routinely explore with these patients [5,10,20]. Informal history taking is currently used to assess mental wellbeing, but this lacks standardisation and can be difficult to interpret. It can also contribute to inequitable care within this patient group. In contrast, the use of objective psychometrics was perceived to be a convenient method to standardise these recommended assessments, especially considering the time constraints of gastroenterology clinicians [29]. However, formal psychometric assessments are not currently used routinely in clinical practice for patients with gastroduodenal DGBIs due to lack of time, lack of knowledge, and wider systemic reasons. Given the indication of acceptance of such psychometrics by both patients and clinicians, psychometrics developed specifically for patients with gastroduodenal DGBIs could therefore be incorporated into external routine medical tests to address these barriers.

These psychometrics could facilitate the consideration of multidisciplinary management plans, which can be tailored to the patient’s individual needs. Both patients and clinicians believed that integrating psychological support into management plans alongside medical treatments would lead to better care and more effective symptom management. Research into multidisciplinary care for patients with DGBIs supports this perspective [30–33]. This could include the use of evidence-based gut-brain psychotherapies, such as cognitive behavioural therapy and gut-directed hypnotherapy, which have reliably shown to be effective at improving the mental health and gastrointestinal symptoms of patients with DGBIs [11,34–36]. These therapies can also be delivered digitally with little effect on efficacy [37–39], which could minimise the concerns raised about the lack of psychologist availability.

Patient stigma was discussed as a potential concern with this approach, an issue which has been reflected in other DGBI research [9,40–43]. Such stigma has been found to have adverse consequences for patients, including non-adherence, reduced treatment effectiveness, lowered healthcare utilisation, and worsened symptoms [41,42,44]. Despite this stigma and fear of being labelled, patients agree that psychology should be integrated as part of their clinical care plan. Therefore, the use of psychometrics should be approached cautiously to reduce stigma and normalise psychometric assessments. For example, the interviewees recommended external handling and integration with medical tests, such as body surface gastric mapping. Previous research has also highlighted the importance of educating patients about the gut-brain axis to reduce patient stigma [20,36,42,43,45]. Communicating to patients in patient-friendly language why this data is being collected and how it will be used and allowing patients to opt out of psychological assessments were reported by the interviewees to be essential in reducing patients’ concerns. Maintaining the therapeutic relationship was also seen as critical to navigating these concerns, with clinicians needing to be mindful to approach psychological assessments in a sensitive, collaborative, patient-centred, and empathetic manner, a finding which has been previously ascertained [15,36,41,45,46]. Training clinicians in these communication skills has been shown to improve patient satisfaction and outcomes [29,47].

Finally, both data corpora identified concerns regarding clinician knowledge of psychological assessments, highlighting the need for improved clinician education. Lack of clinician knowledge has been found to lead to delayed diagnoses, misdiagnoses, and patient frustration [9]. Clinicians should therefore be trained on how to interpret psychometric results and in particular understand when it is necessary to refer to psychologists or multidisciplinary care teams [5,15].

This study achieved thematic saturation and in-depth explorations of the views of both clinicians and patients. However, the study was limited demographically. Although views were gathered from multiple countries, these were limited to English-speaking Western countries and the majority identified as Caucasian. Research has shown that stigma for mental health is higher in Eastern countries [41], and therefore these patients and clinicians may be less receptive to the use of psychometrics. Self-selection bias may have also influenced the results in favour of positive views, as those more receptive and knowledgeable about psychology may have been more likely to volunteer. Future research should therefore explore the views of more diverse populations.

## Conclusions

This study demonstrated that both patients and clinicians see value in integrating psychometrics into routine testing for patients with gastroduodenal DGBIs. Interviews explored how psychometrics could provide useful psychological information to inform a more integrated understanding and approach to symptom management. However, concerns such as patient stigma, lack of clinician knowledge, and poor clinician bedside manner were indicated, especially in relation to their potential consequences for the therapeutic relationship. Despite these concerns, both groups were overall supportive of psychometric integration and provided recommendations about how best to navigate these challenges to ensure psychometrics are used appropriately and effectively in clinical practice. Further research should aim to integrate psychometrics developed specifically for patients with gastroduodenal DGBIs into routine medical tests, with clinician guidance on interpretation. This will allow for the consolidation of physiological and psychological information to aid in the diagnosis and management of patients with gastroduodenal DGBIs.

## Data Availability

All data produced in the present study are available upon reasonable request to the authors

